# Novel Coronavirus (COVID-19) health awareness among the United Arab Emirates Population

**DOI:** 10.1101/2020.09.10.20191890

**Authors:** Balsam Qubais Saeed, Iffat Elbarazi, Mai Barakat

**Affiliations:** Department of Clinical Sciences, College of Medicine, University of Sharjah, P.O. Box 27272, Sharjah, UAE; Sharjah Institute for Medical Research, University of Sharjah, P.O. Box 27272, Sharjah, UAE; Institute of Public Health, United Arab Emirates University, UAE; Department of biochemistry, Faculty of Veterinary Medicine, Mansoura University, Mansoura Egypt

**Keywords:** Knowledge (K), Practices (P), COVID-19, UAE

## Abstract

**Background:** In response to the global (COVID-19) epidemic, the United Arab Emirates (UAE) government is taking precautionary action to mitigate the spread of the virus and protect the safety and well-being of citizens, residents, and visitors. The knowledge and practices of individuals will probably have an important bearing on the course of the coronavirus disease 2019 pandemic. The aim of this study was to evaluate the knowledge and practices toward COVID-19 among the general public in the UAE during the current outbreak COVID-19.

**Methods:** A cross-sectional online survey of 1356 of respondents in the UAE we conducted during the epidemic outbreak between 9^th^ to 24^th^ June-2020. The questionnaire consisted of three sections: Socio-demographic, participants knowledge, and participants practices. Independent-samples t-test, one-way analysis of variance (ANOVA), chi-square, and binary logistic regression have used. A p-value of (p < 0.05) was considered statistically significant.

**Results:** Of the total sample, 72% were females, 47% % were aged between 30-49 years, 57.2% were from Sharjah, 65.6% had a college degree, and 40.6% were unemployed. The total correct score of knowledge and practices questions was high 85% and 90%, respectively.

Males gender, other marital status, and illiterate/primary educational levels had a lower level of knowledge and practices than others. participants aged 18-29 had little higher knowledge than other ages but had a lower level in practices, people who live in Abu Dhabi had better knowledge and practice than other emirates, employed people had a lower level of knowledge but higher in practice. Binary logistic regression analysis presented that females, 18-29 years, and married participants significantly associated with a higher score of knowledge, while female gender, over 30 years old, the martial status of singles, college-level and higher, unemployed, were significantly associated with high mean practice score.

**Conclusions:** To our knowledge, the current study is one of the first studies to evaluate the knowledge and practices of UAE population toward COVID-19. Most of the respondents demonstrate an excellent level of knowledge and awareness as well as proper conscious practices. Continuing to implement the health education programs pursued by the UAE is highly important to maintain the appropriate level of awareness among the public.

## 1. INTRODUCTION

Novel coronavirus (COVID-19), is a respiratory tract infection caused by a new strain of coronavirus, which began in Wuhan, China, in December 2019. However, most patients with uncomplicated COVID-19 have mild symptoms such as fever, cough, headache, muscle pain, sore throat, and nasal congestion, some patients develop severe symptoms that need hospital therapy, about 5% of the patients need admission to an intensive care unit [1]. In some cases, Coronavirus can lead to acute respiratory distress syndrome and organs failure, such as lung, liver, heart, and kidney [1].

The COVID-19 is rapidly spreading from person to person via respiratory droplets, coughing, or sneezing or contaminated hands of people with the http://illness.in addition, the virus can spread through touching a surface with large traces of the virus [2]. Many people get COVID-19 if they breathe in droplets from an infected individual with the virus. That is why it is important to stay two meters (6 feet) away from an infected person. [2]. Based on the epidemiological reports, the incubation period range of COVID-19 virus is 2-14 days, and the virus may be infectious in the patients without symptoms and the infection is more severe in the elderly and people with a chronic health condition[9,10].

Currently, there is no vaccine or effective medication for COVID-19. Implementing infection control measures and public awareness campaigns to limiting the spread of coronavirus infection is the main way to reduce the prevalence of COVID-19 in the community, especially in middle and low-income countries. [3]. In UAE, by11th June of 2020 (when the questionnaire was distributed), there were 41,990 confirmed COVID-19 cases, 26,761 who was fully recovered, 288, deaths while worldwide confirmed cases were 7.41 million and deaths were 418,000 [4]. It is projected that vaccine development will take several months or more, so managing the crisis depends mainly on people’s adherence to recommended health protection measures. These measures are highly influenced by the knowledge and practices of individuals [3].

Many people are taking preventative measures to protect themselves and their families from (COVID-19) all around the world supporting their communities and avoiding the spread of this outbreak. While some people are sharing and apply disappointed information regarding the virus and how to protect against it. Misinformation and an absence of health awareness and information during the Coronavirus outbreak result in individuals not being protected or doing that can harm themselves and others [7].

Public awareness campaigns play an important part in raising the awareness of the public and in drawing their attention to the risks of COVID-19 [22]. The UAE Government handled the situation since the spread of the COVID-19, in March 2020, in a very efficient way and adopted an integrated strategy in maintaining the performance of all sectors in fighting the spread of the disease. Among the measures taken by the UAE government, was distributing a medical guide to health and governmental facilities and the private sector, in addition to other sectors like education, outlets, and tourism. Also, it implemented an intensified awareness campaigns on public hygiene and mandatory sterilization supplies in all places, as well as safe practices to avoid the spread of COVID-19 in public places and in public transport and other public spaces [9].

With the rapid transmission of the COVID-19 virus and the lack of consensus on effective medication or cure for the disease, it is mandatory that the public possess the correct knowledge follow the right practices as a defense line against the disease. Therefore, this study was conducted to evaluate the knowledge and practices toward COVID-19 among the general public in the UAE during the current outbreak COVID-19. The results of the current study are expected to provide baseline data regarding the level of knowledge of individuals and highlight misperceptions and malpractices related to preventive measures hence better planning for effective awareness campaigns and take the appropriate action from local authorities.

## 2. MATERIAL AND METHODS

### 2.1. Study design and participants

This a cross-sectional survey was conducted online and managed by the main author in June-2020. An invitation message using WhatsApp with a link to the survey was sent inviting people to participate and to share this study. The online questionnaire was directed to UAE residents from all nationalities to determine knowledge and practices related to COVID-19 during the current pandemic.

### 2.2. Questionnaire and data collection

A total of 1356 UAE residents participated in this study between 9^th^ -June and 24^th^ -June 2020 from four different emirates (Abu Dhabi, Dubai, Sharjah, and Ajman) in the United Arab Emirates. The survey was created after a thorough search of the literature, Ministry of Health and Prevention (MOHAP) in the UAE and World Health Organization (WHO) reports [10,11,23,24,25]. The survey was reviewed by the research team and by pilot tested by 15 individuals using telephone interviews to clarify and correct any question resulting in a few modifications.

The survey questionnaire consisted of an interface page and three main sections with a total number of 37 questions. The interface of the questionnaire included the title, the objective of the study, information on participants’ confidentiality, and instructions to fill in the survey. The three main parts of the questionnaire included: “socio-demographic data”, their past COVID-19 testing status and COVID-19 test result. The second part on “knowledge regarding COVID-19” consisted of 17 questions includes: Clinical characteristics of virus (K1-K5), the spread of the virus (K6-K9) prevention (K10-K13) and risk factors (K15-K17). The third part of the questionnaire consisted of 11 questions that investigated practices of individuals toward COVID-19 outbreak”. Each questionnaire took approximately10-15 minutes to answer.

### 2.3. Data Analysis

Data analysis conducted using Statistical Package for Social Software (SPSS) version 22. Scale reliability was performed to ensure data consistency, (Cronbach’s alpha coefficient = 0.729) indicating good consistency. Socio-demographic characteristics frequencies, knowledge, and practice answers along with descriptive statistics were presented in mean ± stander deviation SD, while qualitative data were presented in frequency (number\percent). Participants’ knowledge and practice scores were compared with demographics factories using independent-samples t-test, one-way analysis of variance (ANOVA).

To evaluate the knowledge, respondents were given yes, no, and not sure response options each question, A correct response (yes) of each question assigned 1 score, while assigned 0 scores for incorrect respondent (no and not sure). Total knowledge scores ranged from 0-17. Practice response options of always sometimes and never were assigned 2 scores for always, sometimes 1 score, and never 0 scores. Total practice scores ranged from 0-22.

Pearson’s chi-square as appropriate is used to determine the association, correlation, and homogeneity between variables. Binary logistic regression analyses were used to identify factors associated with good, poor knowledge and practices. A p-value of less than 0.05 (< 0.05) was considered statistically significant.

## 3. RESULTS

### Respondents’ Socio-demographic

Out of 1367 received responses, a total of 1356 questionnaire were included in this study, as any incomplete information by participants resulted in an exclusion of their data **Table 1**. Presents the socio-demographic data of the participants. Most of participants were females (72%) and (28%) were males. Of the respondents: (47%) are aged between 30-49 years, (39.8%) are aged between (18-29) years, and (13.2%) were above 50 years. Table 2 shows that (59.7%) are married, (37.6%) are single and 2.7% either divorced or widowed. More than half of respondents are from Sharjah (57.4%), while (19.5%) from Dubai, (15.9) from Ajman, and (7.3%) from Abu Dhabi. With regards to their education level, most participants (65%) had a college degree, while 12%, 21.2% and 1.8% had a postgraduate education, high school/Diploma, and illiterate/primary, respectively. Moreover, 35.8% of participants are employed, 40.6% unemployed, and 23.5% are students. As for the nationality, 19.0% are Emiratis while the rest of the sample (81.0%) are non-Emiratis. Only (18.1%) stated that they were tested for COVID-19 while (81.9%) did not get tested. None of the respondents tested positive for COVID-19, 88.1 and 11.9% included those who had tested negatively and those who stated that they did not know the result of the test, respectively.

**Table 1:**
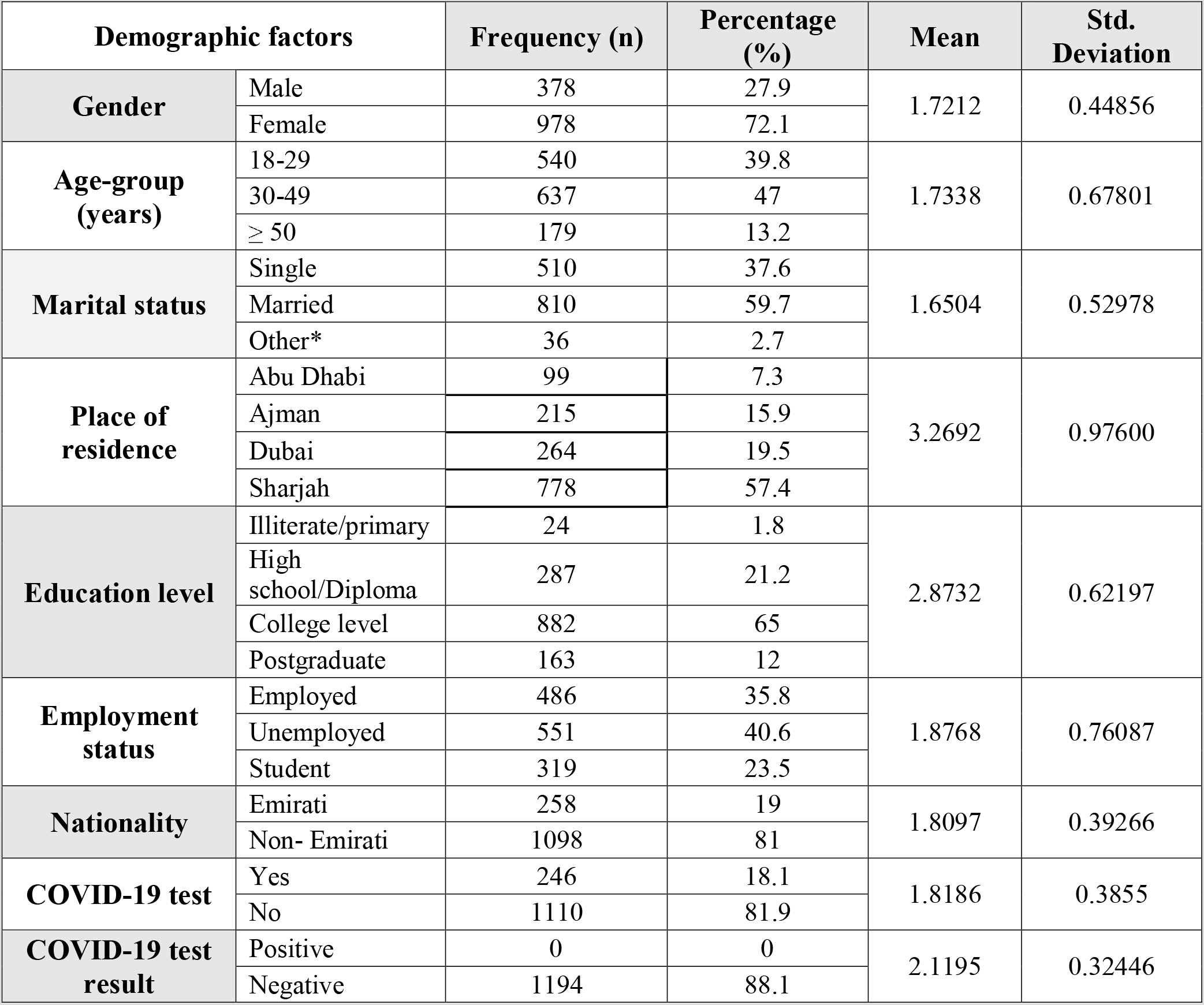
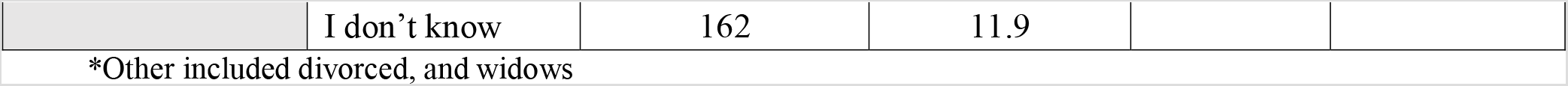
Socio-demographic characteristics of participants, UAE (n = 1356)

| Demographic factors |  | Frequency (n) | Percentage (%) | Mean | Std. Deviation |
| --- | --- | --- | --- | --- | --- |
| Gender | Male | 378 | 27.9 | 1.7212 | 0.44856 |
|  | Female | 978 | 72.1 |  |  |
| Age-group (years) | 18-29 | 540 | 39.8 | 1.7338 | 0.67801 |
|  | 30-49 | 637 | 47 |  |  |
|  | ≥ 50 | 179 | 13.2 |  |  |
| Marital status | Single | 510 | 37.6 | 1.6504 | 0.52978 |
|  | Married | 810 | 59.7 |  |  |
|  | Other* | 36 | 2.7 |  |  |
| Place of residence | Abu Dhabi | 99 | 7.3 | 3.2692 | 0.97600 |
|  | Ajman | 215 | 15.9 |  |  |
|  | Dubai | 264 | 19.5 |  |  |
|  | Sharjah | 778 | 57.4 |  |  |
| Education level | Illiterate/primary | 24 | 1.8 | 2.8732 | 0.62197 |
|  | High school/Diploma | 287 | 21.2 |  |  |
|  | College level | 882 | 65 |  |  |
|  | Postgraduate | 163 | 12 |  |  |
| Employment status | Employed | 486 | 35.8 | 1.8768 | 0.76087 |
|  | Unemployed | 551 | 40.6 |  |  |
|  | Student | 319 | 23.5 |  |  |
| Nationality | Emirati | 258 | 19 | 1.8097 | 0.39266 |
|  | Non- Emirati | 1098 | 81 |  |  |
| COVID-19 test | Yes | 246 | 18.1 | 1.8186 | 0.3855 |
|  | No | 1110 | 81.9 |  |  |
| COVID-19 test result | Positive | 0 | 0 | 2.1195 | 0.32446 |
|  | Negative | 1194 | 88.1 |  |  |
|  | I don't know | 162 | 11.9 |  |  |

\*Other included divorced, and widows

**Table 2:** Descriptive statistics of general knowledge with of participants about COVID-19, UAE (n = 1356)

| Questions |  |  | Yes | No | Not sure | Mean | St. Deviation |
| --- | --- | --- | --- | --- | --- | --- | --- |
| **K1** | COVID-19 is caused by Virus. | N | **1219** | 36 | 101 | 1.17550 | 0.54212 |
|  |  | % | **89.9** | 2.7 | 7.4 |  |  |
| **K2** | Incubation period range of COVID-19 is 2-14 days. | N | **1218** | 36 | 102 | 1.17700 | 0.54436 |
|  |  | % | **89.8** | 2.7 | 7.5 |  |  |
| **K3** | The main clinical symptoms of COVID-19 are fever, tiredness, dry cough, and breathing difficulty | N | **1296** | 24 | 36 | 1.0704 | 0.34407 |
|  |  | % | **95.6** | 1.8 | 2.6 |  |  |
| **K4** | Is there a vaccine for COVID-19? | N | 102 | **1080** | 174 | 2.0521 | 0.44795 |
|  |  | % | 7.5 | **79.6** | 12.8 |  |  |
| **K5** | Is there an active treatment for COVID-19? | N | 168 | **858** | 330 | 2.1167 | 0.59517 |
|  |  | % | 12.4 | **63.3** | 24.3 |  |  |
| **K6** | Are the COVID-19 spreads via respiratory droplets (from coughing, sneezing) of infected people? | N | **1255** | 30 | 71 | 1.1262 | 0.46326 |
|  |  | % | **92.6** | 2.2 | 5.2 |  |  |
| **K7** | Can COVID-19 have transmitted through eyes, in addition to nose and mouth? | N | **1122** | 109 | 125 | 1.2641 | 0.61486 |
|  |  | % | **82.7** | 8.0 | 9.2 |  |  |
| **K8** | Can COVID-19 spreads via through touching contaminated surfaces? | N | **1164** | 78 | 114 | 1.2245 | 0.58449 |
|  |  | % | **85.8** | 5.8 | 8.4 |  |  |
| **K9** | A person with COVID-19 having no fever cannot infect others. | N | 180 | **919** | 257 | 2.0565 | 0.56361 |
|  |  | % | 13.3 | **67.8** | 19.0 |  |  |
| **K10** | Hand washing should be at least 20 minutes. | N | **1110** | 162 | 84 | 3.8230 | 0.71335 |
|  |  | % | **81.9** | 11.9 | 6.2 |  |  |
| **K11.** | We can use hand sanitizer or disinfectant to clean our hands when water is not available. | N | **1254** | 96 | 6 | 1.9336 | 0.26621 |
|  |  | % | **92.5** | 7.1 | 0.4 |  |  |
| **K12** | The minimum distance should you keep it between you and others when go outside is 6 feet (2 meters)? | N | **864** | 132 | 360 | 1.9292 | 0.80643 |
|  |  | % | **63.7** | 9.7 | 26.5 |  |  |
| **K13** | To prevent the spread of COVID-19, individuals should avoid going to crowded places if it's not necessary. | N | **1332** | 0 | 24 | 1.0352 | 0.26314 |
|  |  | % | **98.2** | 0 | 1.8 |  |  |
| **K14** | People who have contact with someone infected with the COVID-19 virus should be immediately isolated in a proper place. | N | **1338** | 6 | 12 | 1.022 | 0.1979 |
|  |  | % | **98.7** | 0.4 | 0.9 |  |  |
| **K15** | It is not necessary for children and young adults to take measures to prevent the infection by the COVID-19. | N | 96 | **1206** | 54 | 1.9677 | 0.3325 |
|  |  | % | 7.1 | **88.9** | 4.0 |  |  |
| **K16** | The virus may be more dangerous in patients | N | **1344** | 0 | 12 | 1.0176 | 0.1869 |
|  | with chronic diseases and elderly. | % | **99.1** | 0 | 0.9 |  |  |
| **K17** | Smokers are likely to be more vulnerable to COVID-19. | N | **1008** | 60 | 288 | 1.4666 | 0.81974 |
|  |  | % | **74.3** | 4.4 | 21.2 |  |  |
| **Total** |  |  | **84.96** |  |  | **1.6151** | **0.4874** |

\* The correct responses for each item are bolded

### Respondents’ knowledge about COVID-19

Table 2. Presents that the prevalence of good knowledge score of the 17 knowledge questions was (1.61 ± 0.48). The correct answers of the questions were presented in table 3 was (85%). Most of the participants (90%) answered that COVID-19 caused by a virus, around (89.8%) answered correctly that the incubation period range of COVID-19 virus is between 2-14 days. Most participants (95.6%) had excellent knowledge about the main symptoms of COVID-19. More than two-thirds of respondence (79.6%) and (63.3%) agreed that no vaccine and treatment is available for COVID-19 until now, respectively. About 92.6%, 82.7% and 85.8% of the participants reported that coronavirus spread through contaminated respiratory droplets of infected people, and that is transmitted through the eyes, in addition to the nose and mouth, and via touching contaminated surfaces, respectively. About (67.8%) reported that the person with COVID-19 having no fever can infect other people. Almost (82%) of participants knew that they should wash their hands at least 20 seconds, while around (92.5%) knew that they can use hand sanitizer if soap is not available. More than half of the participants (63.7%) successfully identified that the minimum distance that should be maintained to prevent the COVID-19 transmission between people is 6 feet (2 meters). Nearly all of the respondents (98.2%) agreed that crowded places should be avoided to reduce virus transmission (98.7%) reported that individuals that contact with person who have COVID-19 positive test should be directly isolated in a suitable place.

**Table 3:** Descriptive statistics of practises of participants towards COVID-19, UAE (n = 1356)

| Questions |  |  | Frequency |  |  |  |  |
| --- | --- | --- | --- | --- | --- | --- | --- |
|  |  |  | Always | Sometimes | Never | Mean | St. Deviation |
| P1 | Do you stay at home and go out only when it is necessary? | N | 1278 | 12 | 66 | 1.1062 | 0.43864 |
|  |  | % | 94.2 | 0.9 | 4.9 |  |  |
| P2 | Have you started to wash or sanitize your hands regularly? | N | 1302 | 12 | 42 | 1.0708 | 0.35753 |
|  |  | % | 96.0 | 0.9 | 3.1 |  |  |
| P3 | Do you wear a mask when you go outside? | N | 1308 | 6 | 42 | 1.0664 | 0.35214 |
|  |  | % | 96.5 | 0.4 | 3.1 |  |  |
| P4 | Do you keep distance between you and other when you go outside? | N | 1278 | 36 | 42 | 1.0885 | 0.37778 |
|  |  | % | 94.2 | 2.7 | 3.1 |  |  |
| P5 | Did you stop going to crowded places recently? | N | 1285 | 42 | 36 | 1.0841 | 0.36083 |
|  |  | % | 94.3 | 3.1 | 2.6 |  |  |
| P6 | Did you stop visiting your relatives and friends during the outbreak? | N | 1200 | 30 | 126 | 1.2080 | 0.59230 |
|  |  | % | 88.5 | 2.2 | 9.3 |  |  |
| P7 | Did you stop kissing your relatives and friends when you meet them? | N | 1296 | 12 | 48 | 1.0796 | 0.37974 |
|  |  | % | 95.6 | 0.9 | 3.5 |  |  |
| P8 | Do you use a credit/debit card or other non-cash modes methods for payment transactions? | N | 960 | 180 | 216 | 1.4513 | 0.75275 |
|  |  | % | 70.8 | 13.3 | 15.9 |  |  |
| P9 | Did you stop sharing your eating utensils and food with others? | N | 1205 | 72 | 79 | 1.1696 | 0.50750 |
|  |  | % | 88.9 | 5.3 | 5.8 |  |  |
| P10 | Did you stop hand shaking with others? | N | 1276 | 8 | 72 | 1.1121 | 0.45374 |
|  |  | % | 94.1 | 0.6 | 5.3 |  |  |
| P11 | Do you follow regular updates on COVID 19? | N | 1065 | 123 | 168 | 1.3385 | 0.68706 |
|  |  | % | 78.5 | 9.1 | 12.4 |  |  |
| Weighted mean |  |  |  |  |  | 1.1613 |  |
| Standard Deviation |  |  |  |  |  | 0.4781 |  |
| Total Main of practices % |  |  |  |  |  | 90.14 |  |

\* The correct responses for each item are bolded

When we asked about the necessity for children and young adults take action to prevent the COVID-19 infection, almost (89%) answered correctly. Almost all participants (99%) answered that the virus is high risk in patients with chronic illness and elderly, and (74.3%) believed that smokers are likely to be more vulnerable to COVID-19 infection. **Respondents’ practices toward COVID-19**

Table 3. Presents participants’ practices toward COVID-19. The prevalence of good practice score among people was (1.16±0.47). The table shows that the total correct answer percentage of the questions on practices of participants to avoid COVID-19 infection was very high (90.14%). Most of the participants (94.2%) answered that they stay at home and go out only when necessary, almost (96%) started to wash or sanitize their hands regularly, and (96.5%) wear a mask when they go outside home. We found that 94.2% of respondents keep a 2-meters distance between them and others when going outside and 94.3% of participants stopped going to crowded places recently. When asked about visits to relatives and friends during the COVID-19 outbreak, 88.5% participants indicated that they stopped visiting them. Majority of participants (95.6%) stopped hugging and kissing their relatives or friends, while 70.8% starting to use a credit/debit card for payment transactions or cashless payment transactions. Our finding showed that 88.9% stopped sharing their food with others and (94.1%) stopped handshaking in greetings with others. Overall, two-thirds of participants (78.5%) follow regular updates on COVID 19 from various information sources.

### Sources of information on COVID-19

Regarding the sources of information of participants on COVID-19, 75.2% indicated that their main source of information were official websites and press releases from the Ministry of Health and Prevention (MOHAP) in the UAE. Among them 42.4% indicated that they were following (WHO) press releases, while 40.7% retrieved their information from different news outlet (Newspaper, Television, Radio) and 50.4% were using social media (Twitter, Facebook, YouTube, WhatsApp, Instagram, and Snapchat). Lastly, 21.2% and 5.75% of participants indicated that their sources of information are mainly from family, friends, and other sources, respectively as displayed in **Table 4** and **Figure 1**.

**Table 4:** Sources of information related to COVID-19 among participants, AE (n = 1356)

| Sources | Participant's response (n) | Participant's response (%) |
| --- | --- | --- |
| Ministry of Health and Prevention-UAE | 1020 | 75.2 |
| WHO press release | 576 | 42.4 |
| News outlet (Newspaper, Television, Radio) | 552 | 40.7 |
| Social media (Twitter, Facebook, YouTube, WhatsApp, Instagram, Snapchat) | 684 | 50.4 |
| Family and friends | 288 | 21.2 |
| Other sources | 78 | 5.75 |

**Figure 1:**
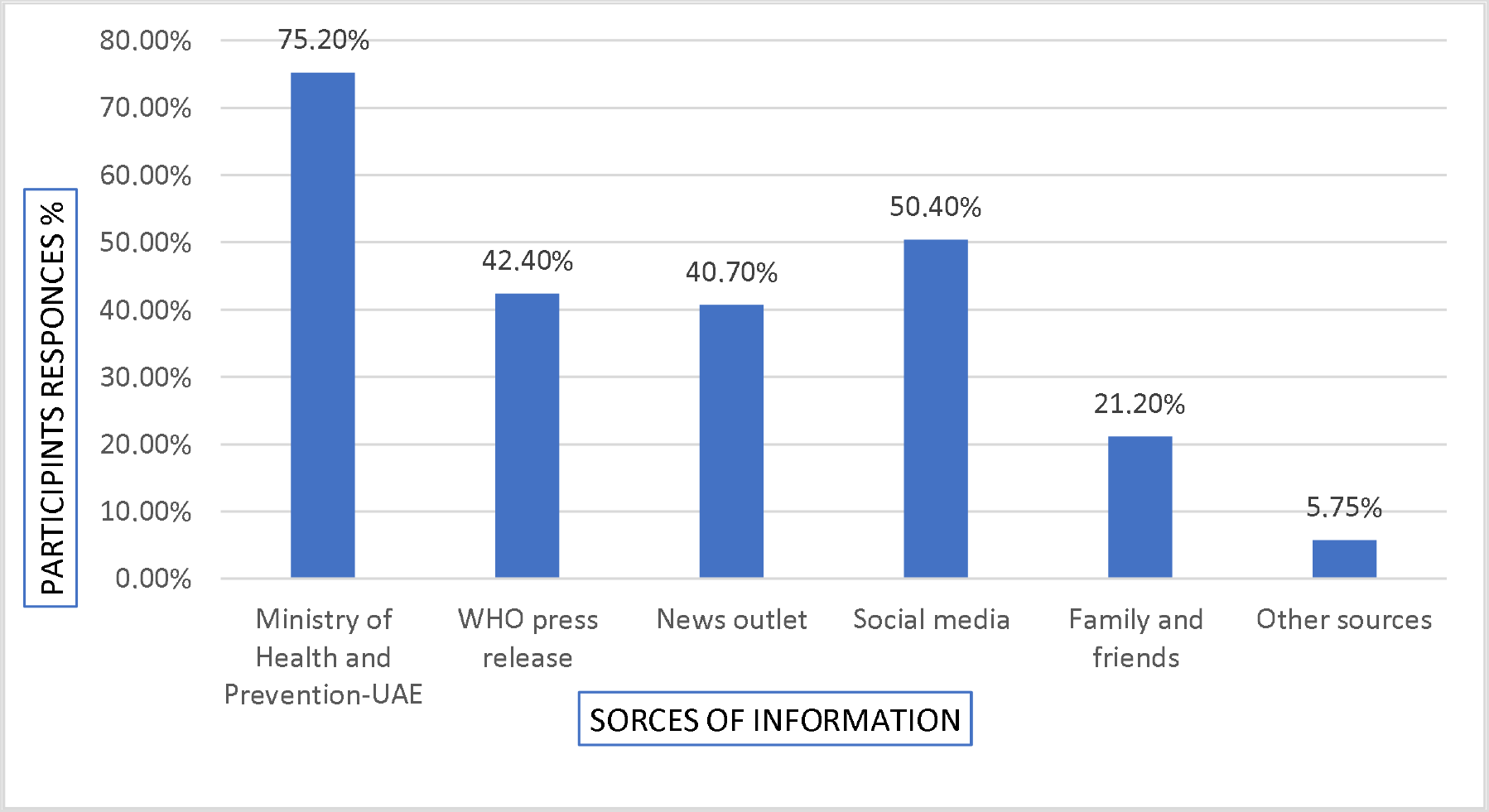
Source of information about COVID-19 among general public in UAE, (n = 1356)

### Correlation between Socio-demographic characteristics with knowledge (K) and practices (P) score of participants, UAE

Table 5. Shows that total scores of knowledge (out of 17) significantly differed across gender, marital status, place of residence, education levels and employment status (p < 0.05), practices scores (out of 11) also significantly differed across of practice with gender, age-groups, marital status, education levels, and employment status (p <0<0.05) while no significant correlation between place of residence and practices (p = 0.077) was identified.

**Table 5:** Socio-demographic characteristics of participants with knowledge (K) and practices (P) score of COVID-19 by demographic variables of participants, UAE (n = 1356)

| Knowledge |  |  |  |  |  |
| --- | --- | --- | --- | --- | --- |
| Demographic variables | | Responses<br>n (%) | Knowledge score<br>(Mean ± SD) | t/f | p-value |
| Gender | Male | 378(27.9%) | 14 ± 2.64 | -8.490 | 0.000* |
|  | Female | 978(72.1%) | **15.09 ± 1.88** |  |  |
| Age-group | 18-29 | 540(39.8%) | **14.94±1.44** | 5.026 | 0.007 |
| | 30-49 | 637(47%) | 14.78± 2.49 | | |
| | ≥ 50 | 179 (13.2%) | 14.35± 2.66 | | |
| Marital status | Single | 510(37.6%) | 14.65±1.97 | 36.074 | 0.000* |
|  | Married | 810(59.7%) | **14.99±2.085** |  |  |
| | Others* | 36 (2.7%) | 12±4.05 | | |
| Place of residence | Abu Dhabi | 99 (7.3%) | **15.33±1.27** | 4.966 | 0.002* |
| | Ajman | 215(15.9%) | 15.10±2.01 | | |
| | Dubai | 264(19.5%) | 14.54±2.13 | | |
|  | Sharjah | 778(57.4%) | 14.71±2.30 |  |  |
| Education level | Illiterate/primary | 24(1.8%) | 9.25±2.32 | 60.928 | 0.000* |
|  | High school/diploma | 287(21.2%) | 14.72±1.83 |  |  |
|  | College level | 882(65 %) | 14.91±2.073 |  |  |
|  | Postgraduate | 163(12%) | 15.04±2.18 |  |  |
| Employment status | Employed | 486(35.8%) | 14.60±2.30 | 5.178 | 0.006 |
|  | Unemployed | 551(40.6%) | 15.01±2.16 |  |  |
|  | Student | 319(23.5%) | 14.67±1.95 |  |  |
| Practices |  |  |  |  |  |
| Demographic Variable |  | Responses<br>n (%) | Practices score<br>(Mean ± SD) | t/f | p-value |
| Gender | Male | 378(27.9%) | 19.29±2.58 | -7.001 | 0.000* |
|  | Female | 978(72.1%) | 20.18±1.87* |  |  |
| Age-group | 18-29 | 540(39.8%) | 19.62 ±2.48 | 12.261 | 0.000* |
|  | 30-49 | 637(47%) | 20.05 ±1.92 |  |  |
|  | ≥ 50 | 179 (13.2%) | 20.45±1.41* |  |  |
| Marital status | Single | 510(37.6%) | 19.74 ±2.46 | 19.422 | <0.000* |
|  | Married | 810(59.7%) | 20.13 ±1.81* |  |  |
|  | Others* | 36 (2.7%) | 18.1 ±2.59 |  |  |
| Place of residence | Abu Dhabi | 99 (7.3%) | 20.47±1.88* | 2.286 | 0.077 |
|  | Ajman | 215(15.9%) | 19.87±2.22 |  |  |
|  | Dubai | 264(19.5%) | 19.87±1.79 |  |  |
|  | Sharjah | 778(57.4%) | 19.90± 2.23 |  |  |
| Education level | Illiterate/primary | 24(1.8%) | 14.75±3.26 | 74.02 | 0.000* |
|  | High school/diploma | 287(21.2%) | 19.33±2.11 |  |  |
|  | College level | 882(65 %) | 20.29±1.88* |  |  |
|  | Postgraduate | 163(12%) | 19.80 ±2.00 |  |  |
| Employment status | Employed | 486(35.8%) | 20.23±1.78* | 14.739 | 0.000* |
|  | Unemployed | 551(40.6%) | 19.98±1.88 |  |  |
|  | Student | 319(23.5%) | 19.41±2.81 |  |  |

\*Other included divorced, and widows
\* The highest score of knowledge and practices are bolded
P: probability \*Significance <0.05

The results score shows that females had a higher level of knowledge (15.09 ± 1.88) and practices (20.18±1.87) than males, people aged of 18-29 years had a very little higher level of knowledge (14.94±1.44) compared with other age groups, while they had lower level in practices (19.62 ±2.48) than other ages. Knowledge (14.99±2.085) and practices (20.13 ±1.81) of married participants were a little higher than those who identified themselves single, widowed and divorced. Residents of Abu Dhabi showed a better knowledge (15.33±1.27) and practices (20.47±1.88) that other emirates. Participants with lower education showed lower level of knowledge (9.25±2.32) and practices (14.75±3.26) than those with higher education while employed participants showed lower knowledge (14.60±2.30) than non-employed, students but shared higher practices with unemployed and students (20.23±1.78) **Table 5**.

### Binary logistic regression associated with the knowledge (K) score (>14.5 and < 14.5) and practices (P) score (>19.5 and < 19.5) as (good and poor) of participants, UAE

Table 6. presents binary logistic regression analysis on variables significantly correlating with participants knowledge and practice toward Coronavirus. The same table presents Odds ratios (ORs) and their 95% confidence intervals (CIs) in a bid to quantify the correlation between factors and the knowledge score (>14 and < 14), and between variables and the practices score (>19 and < 19). The bivariate analysis shows significance when correlated gender, age and marital status and better knowledge. Female gender vs male showed better knowledge (β: 1.146, OR: 3.145, CI:2.235-4.424), age group 18-29 is associated with better knowledge than the age-groups of 30-49 and > 50 respectively (β:-0.917, OR: 0.400, CI: 0.236-0.678), and (β:-1.663, OR:189, CI:0.101-0.355). While being married vs. single participants were significantly associated with the higher mean knowledge (β: 0.559, OR: 1.749, CI:1.017-3.008).

**Table 6:** Binary logistic regression analysis on factors significantly associated with mean knowledge (>14 and < 14) and practice (>19 and < 19) as (good and poor), respectively of COVID-19 of the participants, UAE

|  |
| --- |
| **Knowledge (>14 and < 14)** |

| Variable | $\beta$ | OR (CI 95%) | p-value |
| --- | --- | --- | --- |
| Gender (female vs. male) | 1.146 | 3.145 (2.235-4.424) | 0.000* |
| Age (30-49 vs.>18-29) | -0.917 | 0.400 (0.236-.678) | 0.001* |
| Age ($\geq 50$ vs. 18-29) | -1.663 | 0.189 (0.101-.355) | 0.000* |
| Marital status (married vs. single and others) | 0.559 | 1.749 (1.017-3.008) | 0.043* |
| **Practice (>19 and < 19)** |  |  |  |
| Variable | $\beta$ | OR (CI 95%) | P-value |
| Gender (female vs. male) | 1.619 | 5.050 (3.608-7.069) | 0.000* |
| Age (30-49 vs. 18-29) | 1.134 | 3.110 (2.019-4.790) | 0.000* |
| Age ($\geq 50$ vs. 18-29) | 1.981 | 7.252 (3.893-13.509) | 0.000* |
| Marital status (married vs. single) | -1.149 | 0.317 (0.185-0.543) | 0.000* |
| Marital status (others vs. single) | -2.409 | 0.090 (0.026-0.309) | 0.000* |
| Education level (college level vs. Illiterate/primary) | 2.223 | 9.231(2.238-38.079) | 0.002* |
| Education level (postgraduate vs. illiterate/primary) | 2.022 | 7.553(1.885-30.263) | 0.004* |
| Employment (unemployed vs. employed) | -0.632 | 0.531(0.364-0.774) | 0.001* |
| Employment (students vs. employed) | -1.499 | 0.223(0.132-0.379) | 0.000* |

\*Other included divorced, and widows
P: probability \*Significance <0.05

Moreover, Table 6. presents that gender, age, education and employment status also were significantly associated with good practice while marital status was negatively associated with good practice. Female gender vs male (β: 1.619, OR: 5.050, CI: 3.608-7.069), age group of 30-49 and ≥ 50 had higher practice than score than the youngest group (β: 1.134 OR: 3.110, CI:2.019-4.790), (β: 1.981 OR: 7.252, CI: 3.893-13.509) respectively. Marital status of married and others particularly correlated with poor practice score over the single group (β: –1.149 OR:.317, CI:0.185-0.543) and others (β: –2.409 OR: .0.90, CI:0.026-0.309).

College-level and Postgraduate education (β: 2.223, OR: 9.231, CI:2.238-38.079) and Postgraduate vs Illiterate/Primary (β: 2.022, OR: 7.553, CI:1.885-30.263), were associated with better practices. Being Unemployed or a student showed a poor practice score compare with employed participants (β: –0.632, OR: .531, CI: 0.364-0.774) and (β: –1.499, OR: 0.223, CI:0.132-0.379) respectively.

## 4. DISCUSSION

To our knowledge, this is the first study to explore COVID-19 knowledge and practices toward the COVID-19 pandemic in the UAE and during the peak of the lockdown. Our sample was predominantly young, non-Emiratis, of female gender with middle to higher education levels which represents the UAE population [21]. This also consistent with similar demographics of participants of surveys collected via online platforms. [14]. Our study demonstrated that UAE population and residents have a good to excellent knowledge about COVID-19 causes, risks, precautions, and best practices to avoid infection A similar study conducted in China on COVID-19 Knowledge described the similar trend in respondents in terms of gender and education [9,12] but showed lower knowledge possibly because of the difference in timings related to the spread of COVID-19. Our results showed that almost 85% of our respondents are knowledgeable and aware of the risk, transmission mode, and hygienic and preventive measures nearly matching the same percentage by the same china study and another Jordanian study conducted among medical students and another among the public in Nigeria [9,10,11,12].

We believe that this percentage is high and excellent as knowledge can be explained by two main reasons: the excellent education and action taken by the authorities to contain the COVID-19 spread among the UAE community and /or due to the high level of media coverage including all media outlet and the impact of the pandemic on social life mandating that people read, listen and get educated about it. However, with all the misconceptions that have been spreading around the virus, our respondents had correct information which indicates that the right message is being spread and it could be again explained by the health authorities actions and education campaigns as well respondents’ education level and access to the right information platforms such as (WHO). In contrast to the Jordanian study [11], our results showed that the UAE population may depend more on social media, search engines to get their information about COVID-19.

Our results show similar findings to that of Saudi Arabian study [13] in terms of knowledge, practices, and correlated factors such as gender, age, marital status, place of residents, education level, and employment status as being a mediator for better knowledge and practices. Comparing the socio-cultural context of both countries, the similar findings and results are very reasonable and are considered normal. Moreover, the efforts that the Ministry of health in both countries were highly intensive in terms of testing and in terms of educating the public and to contain the virus which explains the findings and the similarity in results.

Our study has shown that there was better knowledge and better practices associated with younger ages (below 30), gender (Females), education (higher education), and residing in the capital city of UAE (Abu Dhabi) indicating that more education efforts and more intense health education should be directed toward certain populations like men, lower educated levels and people residing outside AD. The higher significance in terms of the emirate can be explained by the fact that more cases were present in Abu Dhabi lately directing authorities to intensify their education campaigns.

Our results showed, when tested through logistic regression analysis, that, gender, age and marital status were significantly correlated with better knowledge a while being married for married was negatively associated with good practice. Being females, with better education level, and being employed were significantly correlated with higher knowledge and correct practices. Our results were compatible with many studies that showed similar significance in terms of better knowledge among the educated. Being a female and a mother is expected to show better knowledge and practices in terms of the COVID-19 precautions and preventions. Better knowledge with higher scores may lead to better practices as shown in this study and as indicated by other studies [11,12,13]. However, our results did not show any significance when we employed the same factors in our regression. We believe that the significance was not really strong due to the fact again of the intensive education by the authorities and the mandatory hygienic and preventive practices such as washing the hands, using sanitizers, wearing masks, keeping distance, and avoiding touching the face [9]. The findings indicate strongly that awareness campaigns especially social media campaigns and strong health promotions approaches and policies that ensure equity through societies can improve people’s adherence to healthy practices and can induce a positive behavioral change no matter what is their educational background, employment or their social status and gender especially in crisis and in sudden public health events [13,14,15] Health literacy is an important factor in improving people’s adherence to precautions, preventions, and screening. However, with the emerging of this pandemic and during crisis management, improving health literacy will depend mainly on people’s access to knowledge, reading, and the ability to reflect critically on complex health issues [16]. Therefore, the efforts on improving awareness and keep spreading the right message are highly mandatory to ensure healthy practices among the community. The COVID-19 crisis is creating a burden on individuals, communities, and authorities that needs a complex integration of efforts to overcome and to manage this crisis. Studies on COVID-19 have pointed strongly to the mental health impacts and the burden the COVID-19 is creating on public health [19,20]. In fact, when asked our participants whether they got tested for COVID-19 18.9% stated that they were tested which indicated again the influence of the UAE government efforts of screening [9]. However, what was not surprising that out of the 11.9% that were not positive some declined to declare their results which indicated that there is a new trend of stigma and stereotype that is emerging with the pandemic spread and that needs to be addressed. In fact, studies have shown that COVID-19 is causing anxiety and affecting mental health due to many factors including the COVID-19 positive result stigma [17,18,19].

## 5. CONCLUSIONS

Overall, UAE residents showed high levels of knowledge and expected practices regarding COVID-19. Similar to most reports around the world, obtaining information depends on official news and information outlets, (WHO), and social media. Strategies to keep the community updated and vigilant about precautions should continue and more focus on specific groups like male and people with limited education.

## Data Availability

all included

## LIMITATIONS

This study was done in a very short time and used convenience samples through an online platform which limits the generalizability of the findings especially that it is expected that people with a fair acceptable level of education will access such surveys. Moreover, knowledge questions are not validated. Another limitation includes the possibility of bias being a cross-sectional study and the possibility of participants to look for answers online.

## DECLARATIONS

### Ethics approval and consent to participate

The study was approved by the Research Ethics Committee (RIC) at University of Sharjah, UAE. The reference number is REC-20-05–31-01, as of 14/06/2020.

### Consent for publication

All authors have revised the manuscript and agreed to the published version of the manuscript in Journal of Health, Population and Nutrition

### Availability of data and material

The data were collected from 9^th^ to 24^th^ June-2020, A total of 1356 UAE residents participated included in this study

### Authors’ contributions

BQ made essential contributions to the conceptualization, study design; methodology; data collection, writing original draft preparation, and review, IE contributed to the writing, review, and editing, MB contributed to the statistical analysis and writing.

## Acknowledgements

The authors thank the University of Sharjah and respondents who participated in the study. Also, they thank Dr. Hellme Abdullah Najim-Family & Community Medicine and Behavioral Sciences department –UOS for review the questionnaire, and for Dr. Rula mudhafar review and modify the statistical analysis of the results.

## List of abbreviations

COVID-19: (Novel Coronavirus)
UAE: (United Arab Emirates)
ANOVA: (analysis of variance)
K: (Knowledge)
P: (Practices)
MOHAP: Ministry of Health and Prevention)
WHO: (World Health Organization)
SPSS: (Statistical Package for Social Software)
SD: (stander deviation)

